# Deep Learning for Detection of Corneal Perforation on Anterior Segment Optical Coherence Tomography in Microbial Keratitis

**DOI:** 10.64898/2026.04.14.26350795

**Authors:** Lucia H. Rohde, Kamini N. Reddy, Folahan Ibukun, Subeesh Kuyyadiyil, Elesh Jain, Gautam Parmar, Rama Chellappa, Nakul S. Shekhawat

## Abstract

**Purpose:** To develop and evaluate deep learning models for automated detection of corneal perforation in microbial keratitis using anterior segment optical coherence tomography (ASOCT) images.

**Methods:** We enrolled 150 patients with microbiologically confirmed keratitis. Contralateral healthy eyes served as controls. Four convolutional neural network models using ResNet architecture were trained and evaluated using ASOCT images to classify the presence or absence of corneal perforation at the eye level. Ground truth labels for perforation were established following consensus grading by two masked ophthalmologist graders. Models differed in inclusion of healthy controls and masking of non-corneal anterior segment anatomy.

**Results:** The best-performing model (Model 1), which included healthy controls and randomly applied masking of the inferior image portion during training, achieved an AUC of 0.965 (95% CI, 0.911-0.995), sensitivity of 84.0% (95% CI, 70.0%-97.1%), and specificity of 97.8% (95% CI, 96.1%-99.3%) for detection of corneal perforation. Models including healthy controls outperformed those without, and lens masking improved discrimination.

**Conclusions:** Deep learning models achieved high diagnostic accuracy for detecting corneal perforation on ASOCT imaging in eyes with microbial keratitis. These findings support the potential role of automated ASOCT analysis as a clinical decision support tool for identifying this vision-threatening complication.

## INTRODUCTION

Microbial keratitis is the leading cause of corneal blindness and the fifth leading cause of blindness globally^[1,2]^. The disease burden is particularly severe in low- and middle-income countries, where agricultural occupations, limited access to eye care, and delayed presentation contribute to poor visual outcomes^[3,4]^. Corneal perforation, which is a full-thickness defect in the corneal stroma caused by inflammatory degradation of stromal collagen fibers in response to severe infection, is associated with profound corneal damage and is one of the most devastating complications of microbial keratitis^[5]^. Left untreated, perforation can lead to prolapse of intraocular contents, endophthalmitis, and loss of the eye^[6,7]^. Management options for perforation range from observation with medical therapy alone for small perforations with iris plugging to procedural interventions for larger perforations such as cyanoacrylate gluing^[8]^, Tenon patch graft^[9,10]^, multilayered amniotic membrane grafting^[11,12]^, or tectonic keratoplasty^[5,13,14]^. Accurate identification of corneal perforation is therefore essential for corneal surgeons to determine the necessity, optimal method, and timing of repair and prevent major adverse outcomes.

Anterior segment optical coherence tomography (ASOCT) has emerged as a clinically useful imaging modality for evaluating corneal infections due to its high-resolution cross-sectional visualization of stromal architecture, infectious infiltrate depth, and deeper anatomic damage that may not be apparent on slit lamp examination alone^[15–21]^. A recent diagnostic concordance study demonstrated that ophthalmologists interpreting ASOCT images were able to detect corneal perforation with substantially greater sensitivity than slit lamp examination, with ASOCT identifying perforation in 16% of eyes compared to 8% via slit lamp biomicroscopy^[21]^. Ophthalmologists interpreting ASOCT images also demonstrated near-perfect inter-grader agreement (κ=0.98) for detection of perforation, indicating that ASOCT could serve as an objective, reproducible, and scalable assessment tool for microbial keratitis. However, ophthalmologist interpretation of ASOCT images requires expertise that is not uniformly available, particularly in remote or resource-limited settings where microbial keratitis is most prevalent.

Deep learning approaches have demonstrated considerable promise across ophthalmic imaging modalities, with successful applications including diabetic retinopathy screening, glaucoma detection, and age-related macular degeneration assessment^[22–24]^. Computer vision algorithms performing automated assessment of ASOCT images could enable rapid, objective, and clinically actionable identification of corneal perforation in microbial keratitis, potentially bridging the expertise gap across varying practice environments. However, the application of deep learning to anterior segment pathology, particularly in the context of microbial keratitis, remains relatively underexplored.

In this study, we developed and evaluated deep learning models for detecting corneal perforation on ASOCT images of eyes with microbial keratitis. We compared model configurations to assess whether inclusion of healthy control eyes in training datasets or masking of non-corneal anatomy affected diagnostic performance. This study adhered to the Standards for Reporting Diagnostic Accuracy Studies for Artificial Intelligence (STARD-AI) guidelines^[25]^.

## MATERIALS AND METHODS

### Study Design and Ethics

This was a cross-sectional diagnostic accuracy study evaluating four deep learning models for detection of corneal perforation on ASOCT imaging. The index test consisted of convolutional neural network-based classification models, and the reference standard was expert consensus grading of perforation status based on multimodal clinical imaging. This study adhered to the tenets of the Declaration of Helsinki and received approval from the Johns Hopkins University School of Medicine institutional review board and the SNC Chitrakoot Eye Hospital institutional research ethics committee. All participants provided written informed consent prior to study enrollment.

### Study Population

We enrolled consecutive patients with microbiologically confirmed bacterial or fungal keratitis treated at SNC Chitrakoot, a tertiary eye care center in Madhya Pradesh, India, between May 2024 and December 2024. Eligible patients had bacterial, fungal, or polymicrobial keratitis confirmed via microbiologic culture or smear microscopy testing. We excluded eyes with ungradable image quality, unreadable ASOCT files, or missing slit lamp examination findings. Contralateral eyes of enrolled patients without evidence of corneal infection, scarring, or perforation were used as healthy controls. Data from baseline visits were included in the algorithm training and evaluation datasets.

### ASOCT Image Acquisition

ASOCT imaging was performed using the Heidelberg Anterion platform (Heidelberg Engineering, Heidelberg, Germany) with the Metrics App, which obtains six radial cross-sectional scans that capture limbus-to-limbus images of the cornea and anterior segment anatomy. This standardized imaging protocol provided comprehensive sampling across multiple evenly spaced meridians of the front of the eye. Additional clinical contextual information used during human grading of ASOCT images included diffuse illumination slit lamp photographs, cobalt blue illumination photographs taken 10-15 seconds after instillation of fluorescein dye, and grayscale photographs of the cornea taken using the Heidelberg Anterion device with a superimposed reference line indicating the exact anatomic cross-section captured on the ASOCT scan **(Supplementary Figure 1).**

### Expert Ground Truth Labeling of ASOCT Images

For each eye, two masked ophthalmologist graders independently evaluated all six radial ASOCT images alongside ancillary imaging and graded the eye for presence or absence of frank corneal perforation. Frank corneal perforation was defined as a full-thickness defect of the stroma with Descemet membrane discontinuity; a collapsed or nearly collapsed anterior chamber along with evidence of severe corneal thinning and/or Descemetocele; and/or overt visualization of iris tissue plugging an area of perforated cornea. Old perforations that had healed with corneal re-epithelialization were not classified as frank perforation. Eyes with iris synechiae extending to the posterior cornea but with a formed anterior chamber and no evidence of severe corneal thinning were also not classified as having active perforation. Following independent grading, disagreements between graders were resolved via a consensus discussion. Inter-grader agreement for ASOCT-based perforation detection was near-perfect (κ=0.98; 95% CI, 0.92-1.00), with only 1 of 150 images (0.7%) requiring adjudication as previously reported^[21]^. Active corneal perforation was defined as an eye-level diagnosis due to its effect on the entire anterior chamber anatomy, and because of the possibility of the six radial cuts failing to capture the exact location of the full-thickness corneal defect.

### Deep Learning Model Development

A deep convolutional neural network was trained to predict the presence of corneal perforation from ASOCT images. Each eye was represented by six radial ASOCT images, and all scans from the same eye were aggregated to produce a single eye-level prediction. A ResNet backbone pretrained on ImageNet was used as the feature extractor. The final fully connected layers were replaced with a custom classification head consisting of a global average pooling layer, dropout regularization, and a sigmoid output layer for binary classification.

Four model variants were trained and evaluated:

1. **Model 1:** Healthy control eyes included; inferior portion of image randomly masked during training to exclude lens anatomy.
2. **Model 2:** Healthy control eyes included; no image masking applied.
3. **Model 3:** No healthy control eyes; no image masking applied.
4. **Model 4:** No healthy control eyes; inferior portion of image randomly masked during training to exclude lens anatomy.

For each eye, one scan was loaded from each angle (0°, 30°, 60°, 90°, 120°, 150°) and concatenated along the channel dimension to create a 6-channel tensor. The inputs were resized to 224x224 pixels and normalized using a mean of 0.5 and standard deviation of 0.25. During training, data augmentation included random horizontal flipping, random rotation and randomly applied masking of the inferior image portion. Models were trained for 20 epochs with a batch size of 16 using the AdamW optimizer with weight decay of 1x10^-4^. Learning rate scheduling was performed using ReduceLROnPlateau based on validation loss. Binary cross entropy with a positive weight of 1.5 times the class imbalance ratio of the training split was used to address class imbalance. Training was performed on a single NVIDIA RTX A5500 GPU.

### Data Partitioning

Data were partitioned at the patient level to prevent data leakage, ensuring that images from the same patient did not appear in both training and test sets. Patient-level stratification was used to preserve the distribution of perforation cases across folds. The dataset was divided into five folds. In each cross-validation iteration, one fold was held out as the test set, one of the remaining folds was used as the validation set for checkpointing and threshold selection, and the remaining three folds were used for training. This process was repeated so that each fold served as the test set once. Model checkpoints within each iteration were selected based on the validation average precision. Final model selection was based on the mean validation F1 score across folds, and reported performance metrics were averaged across the five test folds. **Table 1** summarizes the patient-level split sizes and number of perforation cases in each cross-validation iteration.

**Table 1.** Cross-Validation Splits.

| Iteration | Train Set<br>(Total<br>patients/Positive<br>Perforation) | Validation Set<br>(Total<br>patients/Positive<br>Perforation) | Test Set<br>(Total<br>patients/Positive<br>Perforation) |
| --- | --- | --- | --- |
| 0 | 90/15 | 30/5 | 30/4 |
| 1 | 90/14 | 30/5 | 30/5 |
| 2 | 90/14 | 30/5 | 30/5 |
| 3 | 90/14 | 30/5 | 30/5 |
| 4 | 90/15 | 30/4 | 30/5 |

### Statistical Analysis

Model performance was evaluated using area under the receiver operating characteristic curve (AUC), sensitivity, specificity, F_1_ score, and average precision (area under the precision-recall curve). Ninety-five percent confidence intervals were calculated using bootstrap resampling. Calibration was assessed using calibration curves comparing predicted probabilities to observed outcome frequencies. Optimal classification thresholds were determined using the Youden index in the validation split. Gradient-weighted class activation mapping (Grad-CAM)^[26]^ was used to generate heatmaps visualizing regions of input images that contributed most to model predictions.

## RESULTS

### Study Population

**Table 2** shows demographic and clinical characteristics for the study population, with additional clinical details provided in **Supplementary Table 1**. A total of 150 eyes from 150 patients with microbiologically confirmed microbial keratitis were included in the analysis. The median age was 50.0 years (IQR, 41.0-59.0), and 92 patients (61.3%) were male. Right eyes comprised 64.0% of the study population. The majority of patients (80.0%) presented with visual acuity of logMAR 1.0 or worse, indicating the severity of presenting disease in this high-risk, low-resource rural population. The median duration from symptom onset to presentation was 15.0 days (IQR, 7.0-30.0 days).

**Table 2.** Demographic and Clinical Characteristics of Study Population.

| **Characteristics** | **Value (N=150)** |
| --- | --- |
| **Age, median (IQR)** | **50.0 (41.0-59.0)** |
| **Sex** |  |
| Female, N (%) | 58 (38.7) |
| Male, N (%) | 92 (61.3) |
| **Eye Laterality** |  |
| Right eye, N (%) | 96 (64.0) |
| Left eye, N (%) | 54 (36.0) |
| **LogMAR VA, median (IQR)** | **1.39 (0.6-2.0)** |
| **Perforation Status on ASOCT** |  |
| Present, N (%) | 24 (16.0) |
| Absent, N (%) | 126 (84.0) |
| **Infection Type** |  |
| Fungal Only | 80 (53.3) |
| Bacterial Only | 54 (36.0) |
| Polymicrobial | 16 (10.7) |
| **Stromal Thinning, N (%)** | **46 (30.7)** |
| **Days to Presentation, median (IQR)** | **15.0 (7.0-30.0)** |

Abbreviations: ASOCT = anterior segment optical coherence tomography; IQR = interquartile range; logMAR = logarithm of minimum angle of resolution; N = number; VA = visual acuity

Fungal keratitis was the most common infection type (80 eyes, 53.3%), followed by bacterial keratitis (54 eyes, 36.0%) and polymicrobial infections (16 eyes, 10.7%). Most infiltrates measured 2-6mm in diameter (69.3%), with 26.0% demonstrating posterior one-third stromal involvement. Forty-six eyes (30.7%) had stromal thinning severe enough to be evident on slit lamp examination. Based on consensus ASOCT grading, 24 eyes (16.0%) had frank corneal perforation and 126 eyes (84.0%) did not have perforation. Contralateral healthy control eyes without active corneal infection (N=150) were included in training datasets for Models 1 and 2.

### Model Performance

Model performances are summarized and compared in **Table 3**, with receiver operating characteristic curves, precision-recall curves, calibration curves, and confusion matrices for each model shown in **Figures 1-4**. Model 1 (healthy controls included, masking of deeper anatomy) achieved the best overall discriminative performance, with an AUC of 0.965 (95% CI, 0.911-0.995) and average precision of 0.833 (95% CI, 0.676-0.956). The sensitivity was 84.0% (95% CI, 70.0%-97.1%) and specificity was 97.8% (95% CI, 96.1%-99.3%). The F_1_ score was 79.7% (95% CI, 58.3%-90.3%). Model 2 (healthy controls included, no image masking) achieved an AUC of 0.973 (95% CI, 0.935-0.996), sensitivity of 76.0% (95% CI, 56.5%-92.0%), and specificity of 96.7% (95% CI, 94.7%-98.6%). The F_1_ score was 71.3% (95% CI, 49.0%-82.5%), and the average precision was 0.847 (95% CI, 0.692-0.968). Model 3 (no healthy controls, no image masking) achieved an AUC of 0.944 (95% CI, 0.885-0.983), sensitivity of 64.0% (95% CI, 48.4%-82.7%), specificity of 95.3% (95% CI, 91.8%-98.5%), F_1_ score of 63.5% (95% CI, 40.3%-77.6%), and average precision of 0.821 (95% CI, 0.670-0.942). Model 4 (no healthy controls, masking deeper anatomy) achieved an AUC of 0.941 (95% CI, 0.882-0.984), sensitivity of 76.0% (95% CI, 60.0%-90.0%), specificity of 94.5% (95% CI, 90.3%-97.8%), F_1_ score of 70.1% (95% CI, 49.5%-83.1%), and average precision of 0.795 (95% CI, 0.649-0.938). Across all four model variants, the best overall discriminative performance was observed when healthy controls were included and the inferior portion of the image was masked during training.

**Figure 1.**
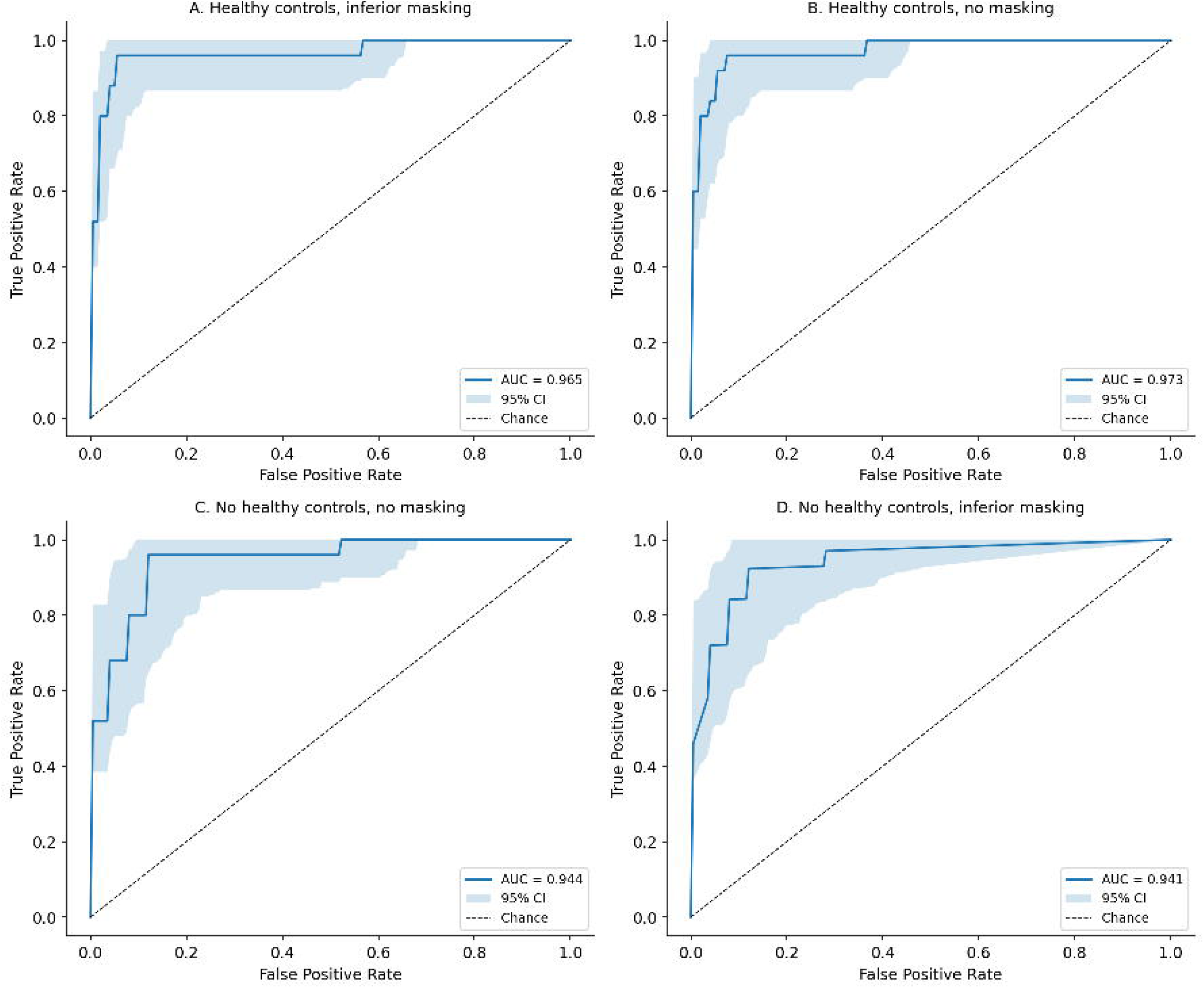
Receiver operating characteristic (ROC) curves for the four deep learning models. (A) Model 1 included healthy controls and applied inferior masking and achieved AUC of 0.965. (B) Model 2 included healthy controls, did not apply masking, and achieved AUC of 0.973. (C) Model 3 did not include healthy controls or masking and achieved AUC of 0.944. (D) Model 4 did not include healthy controls and applied inferior masking and achieved AUC of 0.941.

**Figure 2.**
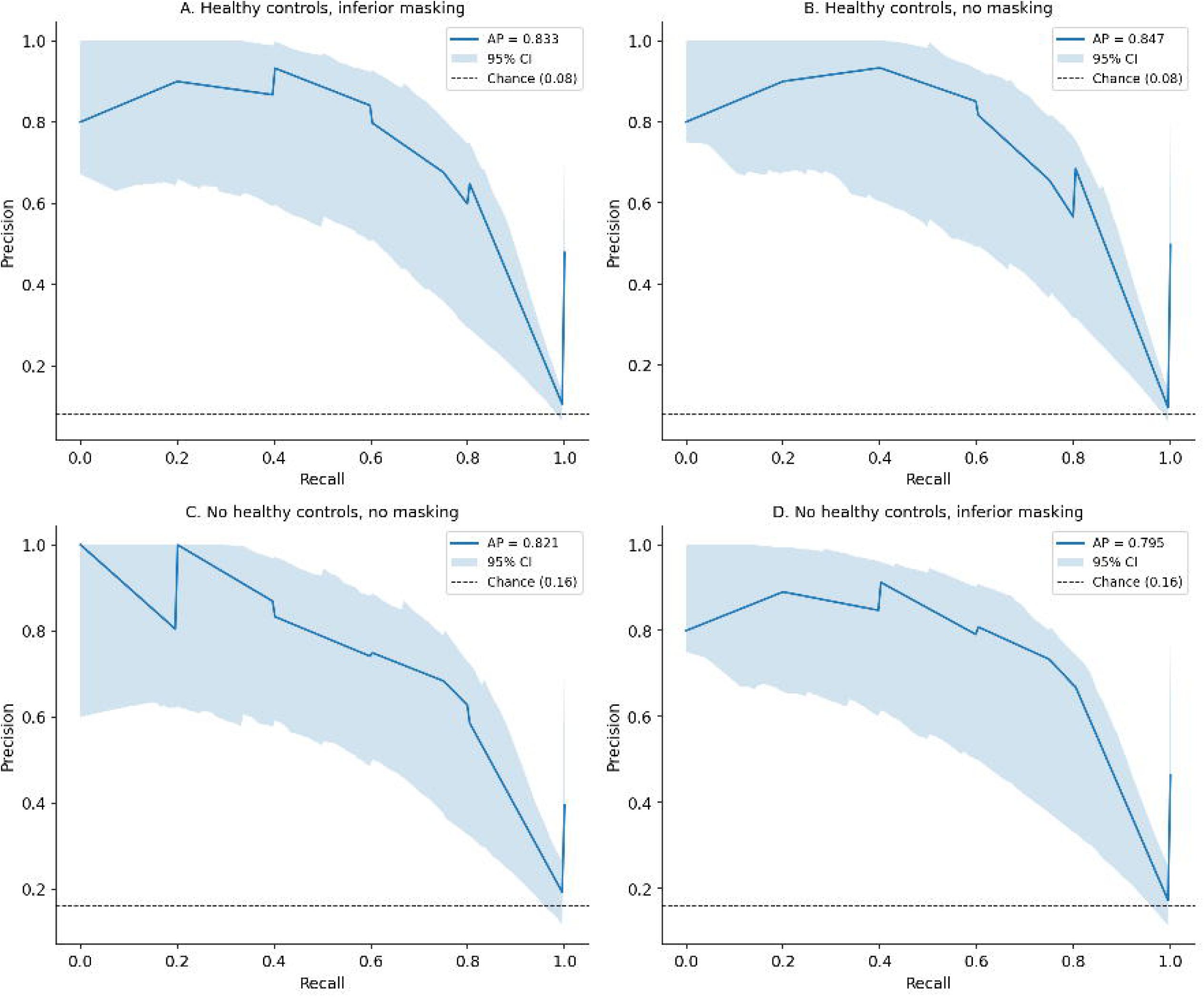
Precision-recall curves for the four deep learning models. The precision-recall curve summarizes the tradeoff between precision and recall across decision thresholds. Model 1 achieved average precision of 0.833, Model 2 achieved 0.847, Model 3 achieved 0.821 and Model 4 achieved 0.795.

**Figure 3.**
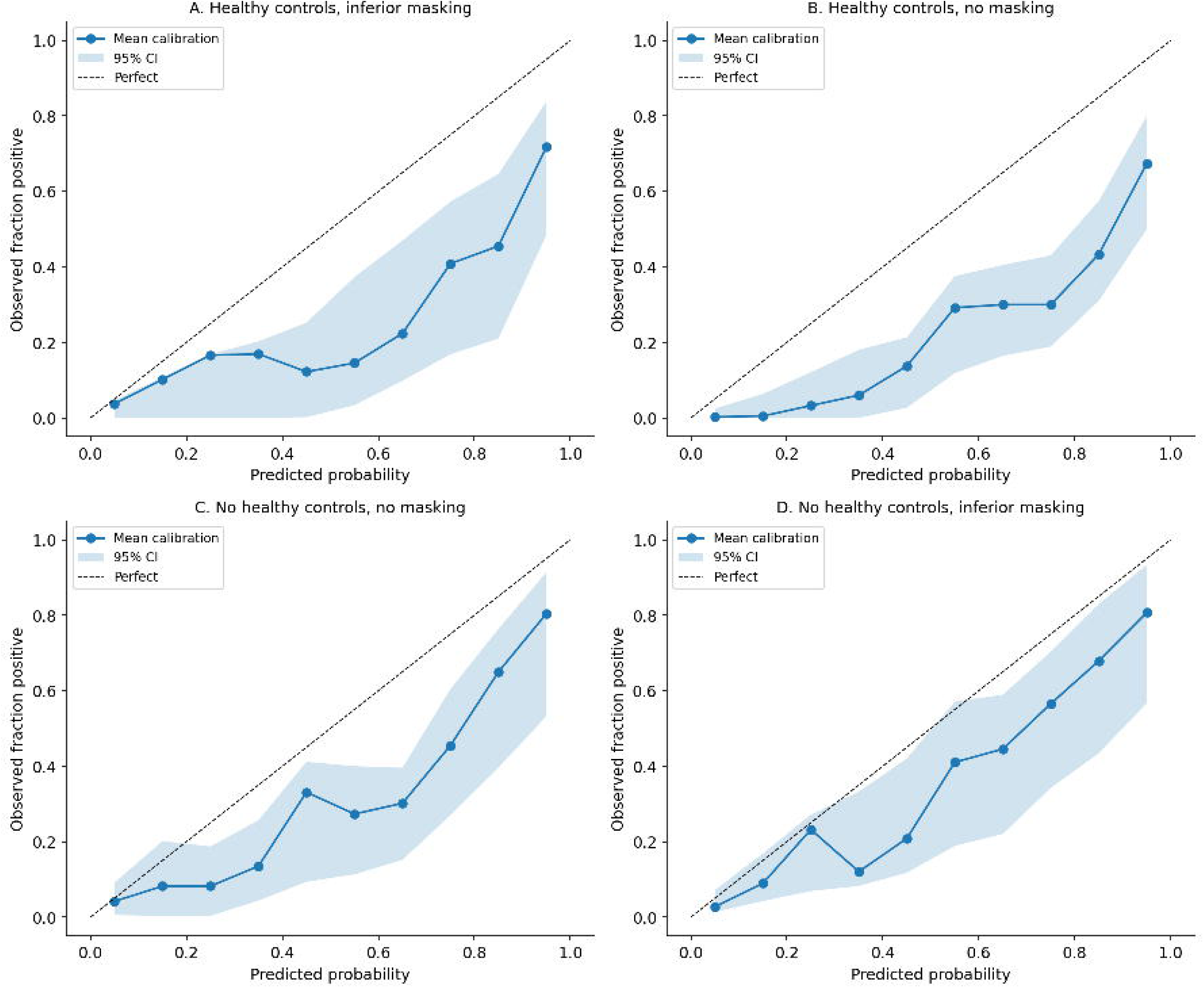
Calibration curves comparing predicted probabilities to observed outcome frequencies. A well-calibrated model produces points close to the diagonal reference line. Model 1 achieved a Brier score and ECE of 0.094 and 0.165, Model 2 achieved 0.118 and 0.202, Model 3 achieved 0.192 and 0.284 and Model 4 achieved 0.092 and 0.136. Although Model 1 achieved the highest discriminative performance, Model 4 demonstrated the highest calibration. Models 2 and 3, which did not include random masking of the inferior portion of the image, demonstrated more overconfidence.

**Figure 4.**
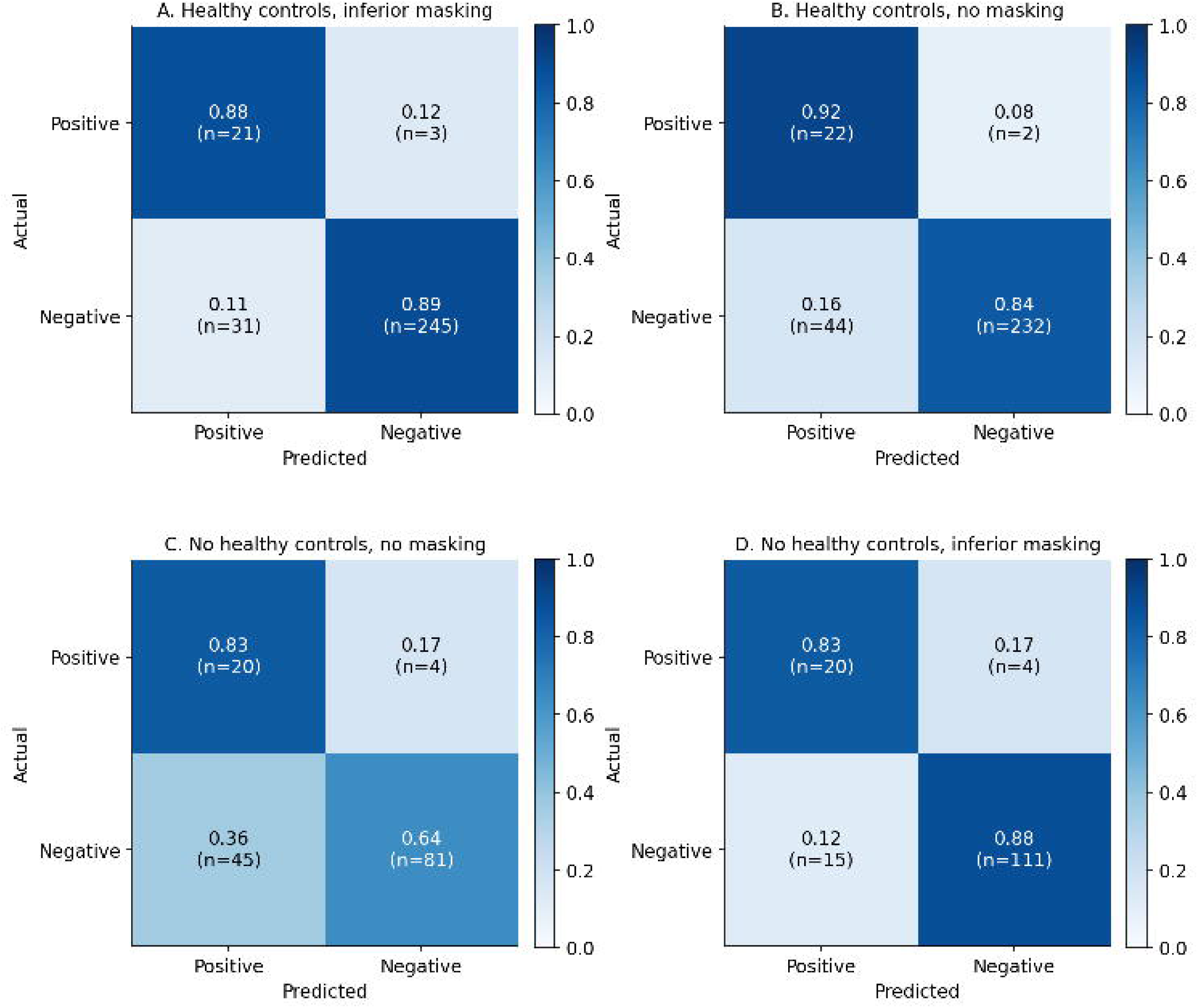
Confusion matrices for the four deep learning models at optimal classification thresholds determined by the Youden index. Model 1 achieved the most favorable balance of sensitivity and specificity, while models trained without healthy controls showed relatively more false negatives.

**Table 3.** Comparison of Deep Learning Model Performance.

|  | **F1 score**<br>**(95% CI)** | **Sensitivity**<br>**(95% CI)** | **Specificity**<br>**(95% CI)** | **ROC AUC**<br>**(95% CI)** | **AP**<br>**(95% CI)** |
| --- | --- | --- | --- | --- | --- |
| **Model 1 (healthy controls, inferior masking)** | 0.797<br>(0.583-0.903) | 0.840<br>(0.700-0.971) | 0.978<br>(0.961-0.993) | 0.965<br>(0.911-0.995) | 0.833<br>(0.676-0.956) |
| **Model 2 (healthy controls, no masking)** | 0.713<br>(0.490-0.825) | 0.760<br>(0.565-0.920) | 0.967<br>(0.947-0.986) | 0.973<br>(0.935-0.996) | 0.847<br>(0.692-0.968) |
| **Model 3 (no healthy controls, no masking)** | 0.635<br>(0.403-0.776) | 0.640<br>(0.484-0.827) | 0.953<br>(0.918-0.985) | 0.944<br>(0.885-0.983) | 0.821<br>(0.670-0.942) |
| **Model 4 (no healthy controls, inferior masking)** | 0.701<br>(0.495-0.831) | 0.760<br>(0.600-0.900) | 0.945<br>(0.903-0.978) | 0.941<br>(0.882-0.984) | 0.795<br>(0.649-0.938) |

Abbreviations: AP = average precision, ROC AUC = area under the receiver operator characteristic curve

### Grad-CAM Visualization

Grad-CAM heatmaps demonstrated distinct patterns of attention across the four models (**Figure 5**; **Supplementary Table 2**). In true positive predictions among eyes with perforation, all four models consistently attended to the anterior chamber, anteriorly displaced iris tissue, and/or areas of iris-cornea touch from collapsed iridocorneal angles, anterior synechiae, or iris plugging of corneal stromal defects. False positives across all models were characterized by attention to iris-cornea touch in eyes with anterior synechiae or shallow anterior chambers without frank perforation. True positives in all four models tended to show minimal activation over deeper non-corneal structures such as the lens. False negatives, though rare, occurred when models attended to structures at or posterior to the lens rather than the anterior chamber. True negative predictions showed the most notable differences in heatmaps across model configurations. Models 1 and 2, which included healthy controls, demonstrated consistent attention to the region at or posterior to the lens. Models 3 and 4, trained without healthy controls, showed more diffuse attention patterns with low spatial specificity, though Model 4 demonstrated somewhat more spatially consistent attention than Model 3.

**Figure 5.**
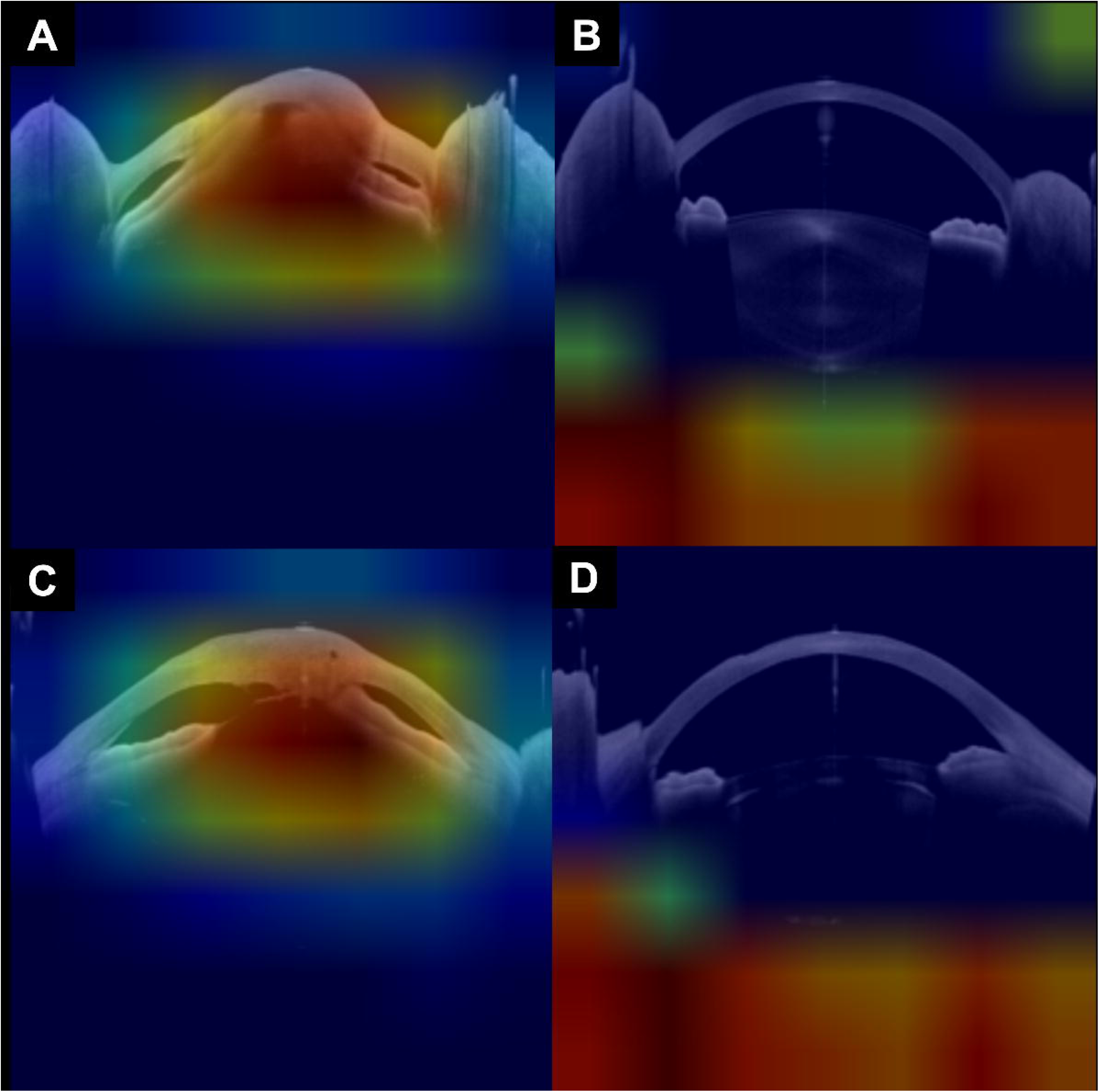
Representative Grad-CAM heatmaps demonstrating patterns of model attention across classification outcomes. (A) Model 1 (healthy controls, inferior masking) true positive: attention focused on anterior chamber collapse and iris-cornea touch in an eye with frank perforation, demonstrating that the model learned clinically relevant anatomic features for perforation detection. (B) Model 2 (healthy controls, no masking) true negative: attention focused on the lens and structures posterior to the lens in a non-perforated eye, reflecting learned association between visibility of deeper anatomy and absence of perforation. This pattern emerged because non-visible lens anatomy occurs more frequently in eyes with severe disease and media opacity, while healthy control eyes have readily visible lens anatomy. (C) Model 3 (no healthy controls, no masking) false positive: attention on iris-cornea touch from anterior synechiae in a non-perforated eye, illustrating the principal failure mode in which the model misclassified eyes with shallow anterior chambers or synechiae that falsely mimic the anatomy of eyes with corneal perforation. (D) Model 4 (no healthy controls, inferior masking) true negative: attention is focused on the inferior region of the image located at or posterior to the lens, despite this region being masked during training. Also observed in Model 1, this pattern’s mechanism remains unclear and warrants further investigation. Of note, Model 3 and 4 exhibited spatially diffuse activation patterns overall, reflecting the absence of healthy control comparators during training, which may have prevented the model from learning to distinguish cornea-specific features from non-specific image characteristics.

Adherence to the STARD reporting guidelines^25^ is summarized in Supplementary Table 3.

## DISCUSSION

This study demonstrates that deep learning models can achieve high diagnostic accuracy for automated detection of corneal perforation on ASOCT imaging of eyes with microbial keratitis. The best-performing model achieved an AUC of 0.965, establishing proof of concept that global eye-level assessments of perforation status can be reliably made using convolutional neural network architectures applied to ASOCT images.

### Impact of Model Configuration on Performance

Our systematic comparison of four model configurations yielded insights relevant to the design of artificial intelligence systems for assessment of anterior segment imaging. First, inclusion of healthy control eyes as negative examples substantially improved model performance. Models trained with healthy controls demonstrated improved sensitivity, potentially because they learned to recognize the full spectrum of normal anterior segment anatomy rather than attempting to distinguish perforated from non-perforated keratitis cases alone. This finding has practical implications for curation of training datasets in future studies.

Second, using randomly applied masking of the inferior image region during training to exclude the inferior portion of images containing deeper anterior segment anatomy such as the lens further improved model discrimination, albeit not by a substantial margin. ASOCT images can contain complex anatomic information of regions beyond the cornea including structural details about the anterior chamber, iris, iridocorneal angle, lens, and in some cases even the anterior vitreous. The lens region alone exhibits substantial variability across eyes due to presence of cataract, pseudophakia, aphakia, or imaging artifacts, none of which are relevant to detecting corneal perforation. By training models to ignore the lens region, we reduced the potential for spurious correlations.

Grad-CAM visualization results provide insight into the differential performance across models (**Figure 5**; **Supplementary Table 2**). All four models attended to clinically meaningful anterior segment structures in true positive predictions, including anterior chamber collapse, anteriorly displaced iris tissue, and iris-cornea touch. This consistency suggests that the models learned anatomic features relevant to detecting perforation regardless of model training configuration. The principal failure mode across all models was false positive classification of eyes with iris-cornea touch (anterior synechiae) or shallow anterior chambers mimicking perforation anatomy, though this occurred infrequently given model specificities ranging from 94.5% to 97.8%.

The most notable differences across model configurations were evident in true negative predictions. For Model 2, which did not employ inferior masking, consistent attention to the region at or posterior to the lens likely reflects learned attention to lens visibility as a discriminative feature, since healthy corneas enable visualization of deeper anatomy whereas opaque or perforated corneas do not. However, lens obscuration can also occur with severe non-perforating conditions such as dense infiltrates or corneal edema, making this an indirect marker that could limit generalizability. Models 3 and 4, trained without healthy controls, showed more diffuse attention in true negatives, consistent with weaker baseline representations of normal anatomy.

Interestingly, both models with inferior masking (Models 1 and 4) demonstrated attention to the region at or posterior to the lens in true negatives, despite this region being masked during training. The mechanism underlying this pattern is unclear. The models may have learned to recognize the masked region itself as an informative feature, or the masking may have influenced how information from adjacent unmasked regions is encoded. Further research using alternative interpretability methods or controlled ablation experiments is needed to clarify how inferior masking affects learned feature representations.

### Clinical Implications

Corneal perforation in the setting of microbial keratitis represents a true ophthalmic emergency, often requiring urgent procedural intervention^[5,14]^. Despite its seriousness, detection of corneal perforation can be challenging when severe corneal infiltrates, scarring, or edema obscure direct visualization of stromal integrity on slit lamp examination. These diagnostic challenges are worse for non-cornea specialists who may lack experience in assessment of corneal infections. Prior work has demonstrated that human graders evaluating ASOCT images can detect substantially more perforations than clinical examination, with slit lamp examination demonstrating only 33.3% sensitivity when ASOCT is used as the reference standard^[15]^. Ophthalmologist graders also show high intra- and inter-grader repeatability for perforation detection on ASOCT images, indicating that such assessments can yield objective and reproducible labels for model training. Assuming ophthalmologist assessments can be replicated by computer vision, an automated system for perforation detection could serve as a scalable clinical decision-support tool to identify high-risk cases needing urgent specialist review, particularly in settings lacking specialist expertise in corneal disease or ASOCT interpretation.

### Implications for Future Models for ASOCT Interpretation

Several implications emerge for the development of future AI systems for anterior segment OCT analysis. First, our improved model performance when training on both diseased and healthy eyes suggests that training datasets should include negative controls, not just diseased eyes without the target condition. Second, deliberate anatomic masking may help focus model attention on clinically relevant regions and reduce model overfitting to spurious correlations. Third, the use of Grad-CAM and similar explainability methods can provide valuable insights into model decision-making that inform iterative model refinement. Future work could explore attention mechanisms or region-of-interest architectures that more explicitly encode anatomical priors. This approach could be extended to other anterior segment structures and conditions detectable on ASOCT images such as hypopyon, endothelial plaque, Descemetocele formation, and iridocorneal lesions. Integration of multimodal imaging data such as slit lamp photographs and/or Scheimpflug tomography may further improve model performance.

### Strengths and Limitations

This study has several strengths. The ground truth labels were established through rigorous masked grading by ophthalmologists who achieved near-perfect inter-grader agreement (κ=0.98), meaning the model was trained using a highly reliable reference standard. Our systematic comparison of four different model configurations enabled mechanistic insights into factors influencing model performance. The use of Grad-CAM visualization produced interpretable evidence supporting our hypotheses about model behavior.

Algorithmic fairness is a key consideration for AI systems intended for clinical deployment^[25]^. Our model was trained on data from a rural population in central India, the country with the highest absolute burden of microbial keratitis worldwide^[27]^. Patients in our dataset presented with clinical characteristics typical of resource-limited settings, including delayed presentation (median 15 days from symptom onset) and a high proportion of fungal infection. This training approach ensures that model development is aligned with the population where AI-assisted perforation detection has substantial potential clinical utility. The anatomic features used by the model to detect perforation, such as full-thickness stromal defects and anterior chamber architecture, are structural findings on ASOCT imaging that are not expected to vary systematically by demographic or geographic factors. Although external validation across heterogeneous clinical settings is required before deployment, training on data from a high-burden population may enhance generalizability to similar settings where the need for automated diagnostic support is high.

Several limitations warrant consideration. First, this study was conducted at a single tertiary referral center in India that serves a rural, low-resource population presenting with severe stages of disease. Although the higher baseline prevalence of perforation in the resulting dataset was ideal for training deep learning models for perforation detection, algorithm performance should be externally validated across heterogeneous geographic, demographic, and clinical settings to verify generalizability prior to clinical deployment. Second, our algorithms were only trained to detect frank perforation, meaning that algorithms were not trained to detect micro-perforations, previously healed perforations, or various forms of profound anatomic damage without perforation such as severe thinning, Descemetoceles, iridocorneal synechiae, or posterior staphylomas. Future research should evaluate algorithm performance for identifying these other clinical features which can also inform clinical and surgical management. Third, because the Heidelberg Anterion ASOCT hardware and imaging settings use specific scan parameters, our results may not be generalizable to other ASOCT devices with different imaging characteristics. Fourth, the relatively small number of perforation cases (n=24) limited statistical power for subgroup analyses and may have affected model calibration. Fifth, we evaluated models using cross-validation rather than a held-out prospective test set, which may overestimate performance compared to real-world deployment. Sixth, this study assessed model accuracy without evaluating the impact of AI assistance on clinical decision-making in real-world settings.

## CONCLUSIONS

Deep learning models achieved high diagnostic accuracy for detecting corneal perforation on ASOCT imaging in patients with microbial keratitis, with the best model achieving an AUC of 0.965. Inclusion of healthy control eyes in the training data and masking of non-corneal anatomy improved model performance. Our findings establish the feasibility of automated ASOCT analysis as a potential clinical decision support tool for identifying this vision-threatening complication of microbial keratitis.

## SUPPLEMENTARY MATERIALS

The following are intended to be included as supplementary materials:

**Supplementary Table 1. Detailed Clinical Characteristics of Study Population**

**Supplementary Table 2. Qualitative Analysis of Grad-CAM Heatmap Patterns Across Model Configurations and Classification Outcomes**

**Supplementary Table 3. STARD-AI Checklist with Manuscript Location References**

**Supplementary Figure 1. Example of ASOCT composite images used for human labeling of perforation in microbial keratitis**

Supplementary Figure 1 legend: Composite images included an ASOCT radial scan (top-left), diffuse illumination slit lamp camera photograph of the eye (top-right), a cobalt blue light slit lamp camera photograph of the eye (bottom-right), and a black-and-white reference thumbnail showing location of radial cut (bottom-left). Each eye had six radial ASOCT scans, each with its own composite image. All six scans were assessed by two expert ophthalmologist graders to achieve an eye-level label perforation versus no perforation. Disagreement between graders was resolved by repeat image review and discussion to achieve consensus. Final consensus labels were used for model training.

## AUTHOR CONTRIBUTIONS

Conceptualization, L.R., R.C. and N.S.; Methodology, L.R., R.C. and N.S.; Software, L.R. and R.C.; Validation, L.R., F.I. and N.S.; Formal Analysis, L.R., K.R. and N.S.; Investigation, L.R., F.I., G.P., R.C. and N.S.; Resources, S.K., E.J., G.P., R.C. and N.S.; Data Curation, L.R., K.R. and N.S.; Writing—Original Draft, L.R. and N.S.; Writing—Review & Editing, L.R., K.R., F.I., S.K., E.J., G.P., R.C. and N.S.; Visualization, L.R. and N.S.; Supervision, S.K., E.J., G.P., R.C. and N.S.; Project Administration, K.R., S.K., E.J., G.P. and N.S.; Funding Acquisition, S.K., E.J., G.P., R.C. and N.S. All authors have read and agreed to the published version of the manuscript.

## IRB STATEMENT

This study was conducted in accordance with the Declaration of Helsinki and approved by the Johns Hopkins University School of Medicine Institutional Review Board (IRB00473135, approved 1/21/2025) as well as the research ethics committee at SNC Chitrakoot.

## INFORMED CONSENT

Informed consent was obtained from all participants involved in the study.

## DATA AVAILABILITY

Datasets and code used in this study may be made available upon reasonable request and in accordance with local regulations. The trained model weights are available at https://github.com/lhrhode/ASOCT_Corneal_Perforation_Detection.

## ACKNOWLEDGMENTS

The authors thank Soumyajit Ray for creating the initial version of composite images. The authors thank the clinical and research staff at SNC Chitrakoot for their assistance with patient recruitment and data collection.

## REFERENCES

1. Bourne R, Steinmetz JD, Flaxman S, et al. Trends in prevalence of blindness and distance and near vision impairment over 30 years: an analysis for the Global Burden of Disease Study. The Lancet Global Health. 2021;9(2):e130–e143. doi:10.1016/S2214-109X(20)30425-3

2. Flaxman SR, Bourne RRA, Resnikoff S, et al. Global causes of blindness and distance vision impairment 1990-2020: a systematic review and meta-analysis. Lancet Glob Health. 2017;5(12):e1221–e1234. doi:10.1016/S2214-109X(17)30393-5

3. Upadhyay MP, Karmacharya PC, Koirala S, et al. Epidemiologic characteristics, predisposing factors, and etiologic diagnosis of corneal ulceration in Nepal. Am J Ophthalmol. 1991;111(1):92–99. doi:10.1016/s0002-9394(14)76903-x

4. Whitcher JP, Srinivasan M, Upadhyay MP. Corneal blindness: a global perspective. Bull World Health Organ. 2001;79(3):214–221.

5. Jhanji V, Young AL, Mehta JS, Sharma N, Agarwal T, Vajpayee RB. Management of corneal perforation. Surv Ophthalmol. 2011;56(6):522–538. doi:10.1016/j.survophthal.2011.06.003

6. Sharma N, Sachdev R, Jhanji V, Titiyal JS, Vajpayee RB. Therapeutic keratoplasty for microbial keratitis. Current Opinion in Ophthalmology. 2010;21(4):293. doi:10.1097/ICU.0b013e32833a8e23

7. Ti SE, Scott JA, Janardhanan P, Tan DTH. Therapeutic keratoplasty for advanced suppurative keratitis. Am J Ophthalmol. 2007;143(5):755–762. doi:10.1016/j.ajo.2007.01.015

8. Rana M, Savant V. A brief review of techniques used to seal corneal perforation using cyanoacrylate tissue adhesive. Cont Lens Anterior Eye. 2013;36(4):156–158. doi:10.1016/j.clae.2013.03.006

9. Kate A, Vyas S, Bafna RK, Sharma N, Basu S. Tenon’s Patch Graft: A Review of Indications, Surgical Technique, Outcomes and Complications. Seminars in Ophthalmology. 2021;0(0):1–9. doi:10.1080/08820538.2021.2017470

10. Shekhawat NS, Kaur B, Edalati A, Abousy M, Eghrari AO. Tenon Patch Graft With Vascularized Conjunctival Flap for Management of Corneal Perforation. Cornea. 2022;41(11):1465–1470. doi:10.1097/ICO.0000000000003068

11. Chan E, Shah AN, O’Brart DPS. “Swiss Roll” Amniotic Membrane Technique for the Management of Corneal Perforations: Cornea. 2011;30(7):838–841. doi:10.1097/ICO.0b013e31820ce80f

12. Grau AE, Durán JA. Treatment of a Large Corneal Perforation With a Multilayer of Amniotic Membrane and TachoSil: Cornea. 2012;31(1):98–100. doi:10.1097/ICO.0b013e31821f28a2

13. Graue-Hernandez EO, Zuñiga-Gonzalez I, Hernandez-Camarena JC, et al. Tectonic DSAEK for the Management of Impending Corneal Perforation. Case Rep Ophthalmol Med. 2012;2012:916528. doi:10.1155/2012/916528

14. Deshmukh R, Stevenson LJ, Vajpayee R. Management of corneal perforations: An update. Indian J Ophthalmol. 2020;68(1):7–14. doi:10.4103/ijo.IJO_1151_19

15. Chong YJ, Azzopardi M, Hussain G, et al. Clinical Applications of Anterior Segment Optical Coherence Tomography: An Updated Review. Diagnostics (Basel). 2024;14(2):122. doi:10.3390/diagnostics14020122

16. Konstantopoulos A, Yadegarfar G, Fievez M, Anderson DF, Hossain P. In vivo quantification of bacterial keratitis with optical coherence tomography. Invest Ophthalmol Vis Sci. 2011;52(2):1093–1097. doi:10.1167/iovs.10-6067

17. Konstantopoulos A, Kuo J, Anderson D, Hossain P. Assessment of the Use of Anterior Segment Optical Coherence Tomography in Microbial Keratitis. American Journal of Ophthalmology. 2008;146(4):534–542.e2. doi:10.1016/j.ajo.2008.05.030

18. Soliman W, Fathalla AM, El-Sebaity DM, Al-Hussaini AK. Spectral domain anterior segment optical coherence tomography in microbial keratitis. Graefes Arch Clin Exp Ophthalmol. 2013;251(2):549–553. doi:10.1007/s00417-012-2086-5

19. Abdelghany AA, Alio JL, AttaAllah HR. Role of Anterior Segment Optical Coherence Tomography in Staging and Evaluation of Treatment Response in Infectious Keratitis. Cornea. 2024;43(10):1216–1222. doi:10.1097/ICO.0000000000003466

20. Khalil H, Bolz M, Waser K, et al. Diagnostic Potential of Anterior Segment Optical Coherence Tomography Scans for Pseudomonas Keratitis. Translational Vision Science & Technology. 2023;12(11):34. doi:10.1167/tvst.12.11.34

21. Ibukun F, Reddy K, Kuyyadiyil S, Jain E, Parmar G, Shekhawat NS. Detection of Infectious Corneal Perforation Using Anterior Segment Optical Coherence Tomography. medRxiv. Published online January 30, 2026:2026.01.28.26345085. doi:10.64898/2026.01.28.26345085

22. De Fauw J, Ledsam JR, Romera-Paredes B, et al. Clinically applicable deep learning for diagnosis and referral in retinal disease. Nat Med. 2018;24(9):1342–1350. doi:10.1038/s41591-018-0107-6

23. Gulshan V, Peng L, Coram M, et al. Development and Validation of a Deep Learning Algorithm for Detection of Diabetic Retinopathy in Retinal Fundus Photographs. JAMA. 2016;316(22):2402–2410. doi:10.1001/jama.2016.17216

24. Ting DSW, Cheung CYL, Lim G, et al. Development and Validation of a Deep Learning System for Diabetic Retinopathy and Related Eye Diseases Using Retinal Images From Multiethnic Populations With Diabetes. JAMA. 2017;318(22):2211–2223. doi:10.1001/jama.2017.18152

25. Sounderajah V, Guni A, Liu X, et al. The STARD-AI reporting guideline for diagnostic accuracy studies using artificial intelligence. Nat Med. 2025;31(10):3283–3289. doi:10.1038/s41591-025-03953-8

26. Selvaraju RR, Cogswell M, Das A, Vedantam R, Parikh D, Batra D. Grad-CAM: Visual Explanations from Deep Networks via Gradient-Based Localization. Int J Comput Vis. 2020;128(2):336–359. doi:10.1007/s11263-019-01228-7

27. Ung L, Bispo PJM, Shanbhag SS, Gilmore MS, Chodosh J. The persistent dilemma of microbial keratitis: Global burden, diagnosis, and antimicrobial resistance. Survey of Ophthalmology. 2019;64(3):255–271. doi:10.1016/j.survophthal.2018.12.003

