## Supplementary figures and images for "Deep Learning for Detection of Corneal Perforation on Anterior Segment Optical Coherence Tomography in Microbial Keratitis"

### Supplementary figure1

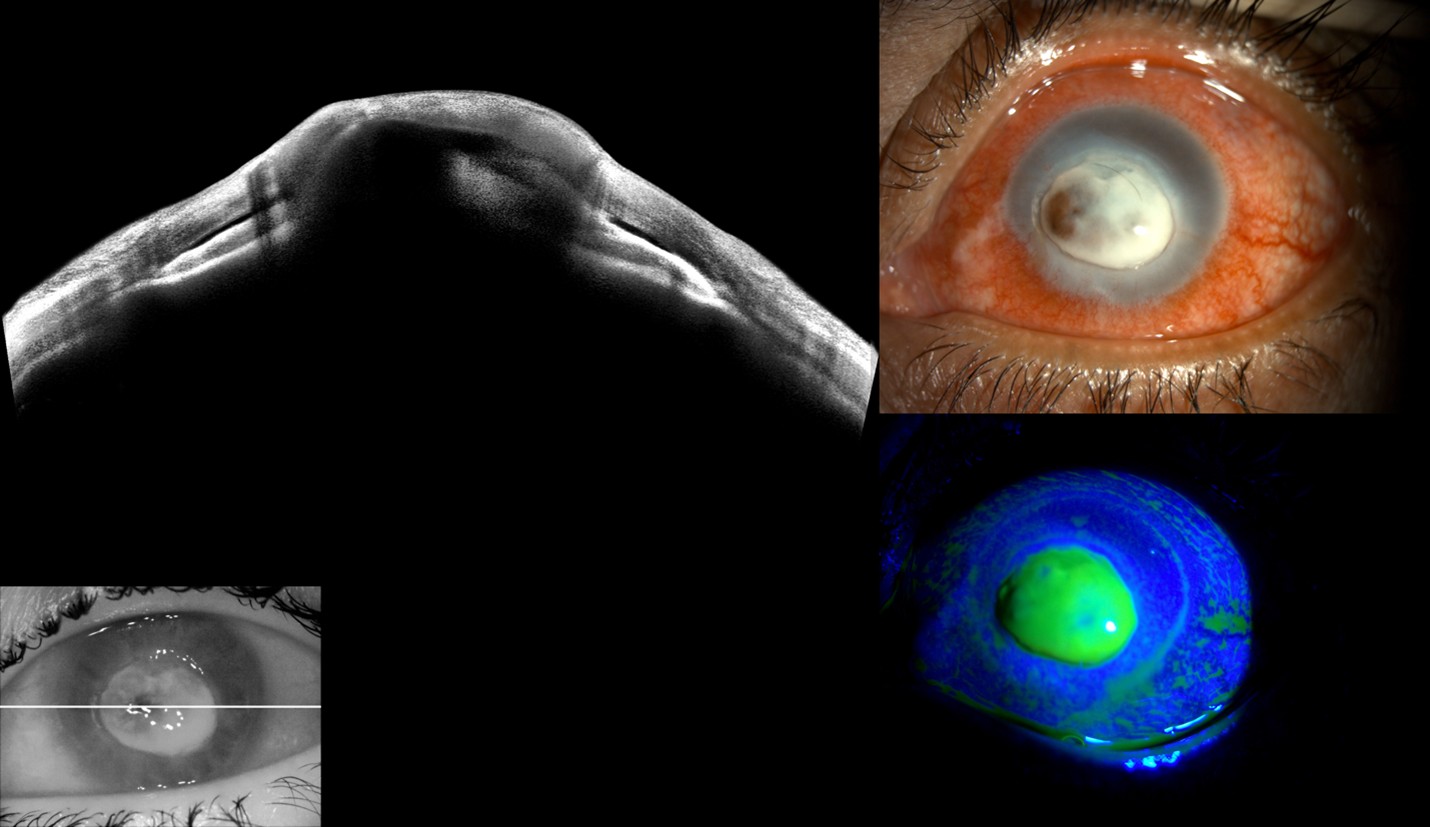
