## Supplementary Table 1-3 for "Deep Learning for Detection of Corneal Perforation on Anterior Segment Optical Coherence Tomography in Microbial Keratitis"

**Supplementary Table 1. Detailed Clinical Characteristics of Study Population**

| **Characteristics** | **Value (Total N=150)** |
| --- | --- |
| **Snellen visual acuity, N (%)** |  |
| 6/6 to ≤6/18 | 17 (11.3) |
| 6/24 to ≤6/36 | 13 (8.7) |
| 1/60 to ≤6/60 | 32 (21.3) |
| Finger counts or Hand movements | 36 (24.0) |
| Light perception or No light perception | 52 (34.7) |
| **Lens Status, N (%)** |  |
| Clear crystalline lens | 57 (38.0) |
| Early lens changes | 31 (20.7) |
| Immature cataract | 31 (20.7) |
| Pseudophakia | 8 (5.3) |
| Unable to determine | 22 (14.7) |
| **Infiltrate Diameter** |  |
| 0 to <2mm | 21 (14.0) |
| 2 to <6mm | 104 (69.3) |
| ≥6mm | 24 (16.0) |
| **Infiltrate Depth** |  |
| Anterior 1/3 Stroma | 133 (88.7) |
| Middle 1/3 Stroma | 97 (64.7) |
| Posterior 1/3 Stroma | 39 (26.0) |
| **Infiltrate Within 2mm of Limbus** | 13 (8.7) |

Abbreviations: mm = millimeter; N = number

**Supplementary Table 2. Qualitative Analysis of Grad-CAM Heatmap Patterns Across Model Configurations and Classification Outcomes**

| **Model** | **Outcome** | **Primary Anatomic Region of Attention** | **Characteristic Attention Pattern** | **Clinical Interpretation** |
| --- | --- | --- | --- | --- |
| Model 1 (healthy controls, inferior masking) | True positive | Anterior chamber | Anterior chamber highlighted in nearly all true positive eyes; lens anteriorly displaced into position where anterior chamber would normally be located; broad iris-cornea touch regions activated | Model correctly identified anterior chamber collapse and iris displacement as markers of perforation |
|  | True negative | At or posterior to lens | Nearly all eyes show attention over a horizontal region at or posterior to lens which is consistent with inferior masked region; occasional attention to abnormal iris anatomy in infected eyes | Model's consistent attention to inferior masked regions may reflect recognition of mask itself as an informative feature, or encoding of information from adjacent unmasked regions |
|  | False positive | Anterior chamber | All false positives show attention over areas of iris-cornea touch and/or shallow anterior chambers without frank perforation | False positives occurred in eyes with anterior synechiae mimicking perforation anatomy |
|  | False negative | Posterior to cornea | Attention focused on horizontal region posterior to cornea, more so than anterior chamber | Missed perforations occurred when model attended to deeper structures located at or posterior to the lens rather than within the anterior chamber |
| Model 2 (healthy controls, no masking) | True positive | Anterior chamber and lens region | Anterior chamber highlighted in all true positive eyes; anteriorly displaced lens and iris-cornea touch areas also activated | Similar detection pattern to Model 1 |
|  | True negative | At or posterior to lens | Nearly all eyes show attention over a horizontal region at or posterior to lens; some attention to iris-cornea touch or anteriorly bowed iris | Model learned that visibility of the lens and structures posterior to it indicates a non-perforated eye; healthy controls establish baseline representation of normal anatomy |
|  | False positive | Iris-cornea interface | Attention focused on iris-cornea touch, with iris abnormalities and attention typically more prominent on one side of anterior chamber | Similar error mode to Model 1; anterior synechiae without frank perforation triggered false positive classification |
|  | False negative | Lens region | Attention over region posterior to anteriorly displaced iris, in presumed location of lens | Model attended to lens region rather than recognizing anterior chamber collapse |
| Model 3 (no healthy controls, no masking) | True positive | Anterior chamber and iris-cornea touch | Attention focused on anterior chamber region posterior to anteriorly bowed iris, or directly over areas of iris-cornea touch | Correct anatomic attention, but model lacked baseline representation of normal anatomy |
|  | True negative | Variable and diffuse | Less consistent pattern; many eyes show attention over region posterior to lens; some show attention over anterior chamber or areas of iris abnormality | Without healthy controls, model developed weaker representation of normal anatomy; attention patterns less spatially organized |
|  | False positive | Posterior to iris | Attention over area posterior to anteriorly bowed iris | Similar error pattern to other models |
|  | False negative | Iris abnormalities | Iris-cornea touch and iris abnormality highlighted | Model attended to relevant structures but failed to classify correctly |
| Model 4 (no healthy controls, inferior masking) | True positive | Anterior chamber | All true positive eyes show attention over anterior chamber; some eyes also show attention to areas of focal or broad iris-cornea touch occurring due to collapsed anterior chamber | Correct anatomic attention; inferior masking may have focused attention on anterior structures |
|  | True negative | Posterior portion of image | Most eyes show attention over region posterior to lens; several eyes show attention over areas of focal iris abnormality such as synechiae or anteriorly bowed iris | More consistent attention than Model 3, suggesting inferior masking provided some regularization; however, attention to masked region despite its absence during training requires further investigation |
|  | False positive | Variable | Variable attention over iris-cornea touch, lens region posterior to bowed iris, or anterior chamber | Heterogeneous error patterns without single failure mode of mis-identification |
|  | False negative | Anterior chamber | Anterior chamber with focal iris-cornea touch highlighted | Model attended to correct anatomic region but still misclassified |

**Supplementary Table 3. STARD-AI Checklist with Manuscript Location References**

| **TITLE/ABSTRACT** | | |
| --- | --- | --- |
| 1 | Identify as AI diagnostic accuracy study | Title, Abstract |
| 2 | Structured summary | Abstract |
| **INTRODUCTION** | | |
| 3 | Scientific/clinical background, intended use | Introduction |
| 4 | Study objectives and hypotheses | Introduction, para 4 |
| **METHODS** | | |
| 5 | Study design | Methods: Study Design |
| 6 | Ethics approval and consent | Methods: Ethics |
| 7 | Eligibility criteria | Methods: Study Population |
| 11 | Data source and collection | Methods: ASOCT Acquisition |
| 12 | Dataset annotation | Methods: Reference Standard |
| 13 | Devices/software for index test | Methods: Model Development |
| 14 | Data acquisition protocols | Methods: ASOCT Acquisition |
| 15b | Dataset partitioning | Methods: Data Partitioning |
| 19 | Reference standard definition | Methods: Reference Standard |
| 20 | Rationale for reference standard | Methods: Reference Standard |
| 21-22 | Statistical methods | Methods: Statistical Analysis |
| 23 | Algorithmic bias methods | Patient-level partitioning to prevent leakage and model evaluation across cross-validation folds with stratification for perforation status. |
| **RESULTS** | | |
| 24 | Flow of participants | Results: Study Population |
| 25 | Test set characteristics | Results: Study Population |
| 26-27 | Cross-tabulation and accuracy | Results: Model Performance, Table 3 |
| 28 | Test set representativeness | Results: Study Population |
| 29 | Performance error analysis | Results: Grad-CAM |
| **DISCUSSION** | | |
| 33-34 | Limitations, implications | Discussion: Strengths/Limitations |
| 35 | Fairness considerations | Discussion: Strengths/Limitations |
| **OTHER** | | |
| 36 | Study registration | N/A |
| 37-38 | Protocol, funding | Funding section |
| 39 | Commercial interests | Disclosures |
| 40a-b | Data/code availability, audit | Data Availability |

This checklist follows the STARD-AI reporting guideline (Sounderajah V, et al. Nat Med. 2025;31:3283-3289).
